# Microbiological and Clinical Characteristics of Hypervirulent *Klebsiella pneumoniae* isolated from Patients in Tertiary Centers

**DOI:** 10.1101/2023.11.17.23298703

**Authors:** Aimi Khairuddin, Nik Mohd Noor Nik Zuraina, Nur Syafiqah Mohamad Nasir, Chua Wei Chuan, Hamimi Salihah Abdul Halim, Wardah Yusof, Azura Husin, Chan Yean Yean, Siti Asma’ Hassan

**Author notes:** **Corresponding author:** Associate Prof. Dr Siti Asma’ Hassan. These authors contributed equally to this work and shared the first authorship (Aimi Khairuddin, Nik Mohd Noor Nik Zuraina).

## Abstract

Hypervirulent *Klebsiella pneumoniae* (hvKp) has emerged as a significant pathogen capable of causing severe community-acquired infections in otherwise healthy people. This cross-sectional, retrospective study aimed to investigate the prevalence of hvKp, its virulence-associated genes, and the clinical manifestations of hvKp infections. HvKp was defined in this study as *K. pneumoniae* with a positive string test and harbouring the serotype K1 or K2 gene. A total of 180 isolates from various clinical specimens were collected from June 2020 to June 2021 in four main hospitals in Kelantan. All isolates were examined for hypermucoviscous phenotype by string test, while the presence of capsular serotype and other virulence genes (*rmpA, rmpA2, iucA, magA, peg-344*) was done by PCR. Patients’ clinical data was collected and analyzed. String test positive isolates (23.8%, n = 43) were identified as hypermucoviscous *K. pneumoniae* (hmKp). Capsular serotypes K1 and K2 were detected in 11.1% (n = 20) and 6.1% (n = 11), respectively. The prevalence of hvKp was found to be 9.4% (n = 17). All the hvKp isolates were positive for *rmpA, rmpA2, iucA,* and *peg-344* genes, while all ten hvKp-K1 serotypes were positive for *magA*, the K1serotype-specific gene. The associations of all the corresponding virulence genes with both serotypes K1 and K2 were statistically significant (p<0.05). HvKp infections were more prevalent in men and individuals with hypertension. Pneumonia was the leading clinical diagnosis in hvKp infected patients, with the mortality rate was 12%. The presence of all biomarkers (*rmpA, rmpA2, magA* (for K1 serotype), *iucA,* and *peg-344* in hmKp, in combination with clinical manifestations, might be reliable for hvKp diagnosis and epidemiological surveillance.

## Introduction

*Klebsiella pneumoniae* can be classified as either classical *K. pneumoniae* (cKp) or hypervirulent *K. pneumoniae* (hvKp) strains based on their phenotypic and clinical characteristics. In contrast to cKp, hvKp typically exhibits a hypermucoviscous phenotype via a string test, in which a viscous string of at least five millimetres can be stretched from the colony using an inoculation loop. In addition, hvKp has the ability to infect healthy individuals of any age, as well as the proclivity to cause multiple sites of infection and/or metastatic spread in the community.

Despite being hypermucoviscous, hvKp strains employ distinct virulence determinants such as siderophores, excessive capsule production, lipopolysaccharides, and colibactin toxin for survival and pathogenesis. The polysaccharide capsule is believed to be the key virulence factor responsible for distinctive hypermucoviscous characteristic [1]. Capsular serotypes K1 and K2 have been reported as the most common strains associated with hvKp infections. The ability to produce enhanced amount of capsular polysaccharides is mediated by *rmp*A and *rmp*A2 hvKp-specific regulators of the mucoid phenotype [2, 3]. The *mag*A gene, which is recognized as a K1 surrogate marker, encodes a polymerase involved in capsule synthesis [4]. HvKp strains produce four different plasmid-encoded siderophores (aerobactin, enterobactin, salmochelin, and yersiniabactin) for iron acquisition. Aerobactin, encoded by the *iuc* gene, is expressed in more than 90% of hvKp strains but only 6% of cKp strains. Hence, it is suggested that aerobactin can serve as a more reliable biomarker for hvKp [5].

The prevalence of hvKp varied among studies. The first clinical report published in 1986 described seven cases of invasive *K. pneumoniae* infection in Taiwanese community members who presented with a hepatic abscess in the absence of biliary tract illness and septic endophthalmitis [6]. *K. pneumoniae* strains causing hepatic abscesses in the Taiwanese patients had a higher likelihood of being hypermucoviscous than non-invasive strains. Lin *et al.* (2012) reported that healthy subjects from Asian countries, including Malaysia, had a higher prevalence of *K. pneumoniae* colonic colonization (18.8% to 87.7%), in contrast to Western countries (5% to 35%) [7]. However, data on the prevalence of hvKp is limited. Therefore, this study aimed to determine the prevalence of hvKp and its associated virulence genes, as well as to describe the clinical presentations of patients infected with this pathogen. In this study, hvKp is defined as *K. pneumoniae* isolates having a hypermucoviscous phenotype and the presence of either the KI or K2 gene.

## Methods

### Study design

This cross-sectional study involved retrospective record reviews from June 2020 until June 2021. Clinical isolates from various types of specimens were collected from four tertiary reference hospitals in Kelantan, Malaysia. The inclusion criteria for this study included *K. pneumoniae* isolates that were confirmed by both culture and molecular methods, via routine diagnostic tests and via PCR detection of the housekeeping gene (*gap*A), respectively. *K. pneumoniae* isolates from the same patient with the same episode of infection were excluded. A total of 180 isolates from various clinical specimens were collected during the study period and subjected to string test and other molecular methods. Operational definitions used in this study were: a) hmKp - a hypermucoviscous *K. pneumoniae* colony with positive string test (defined by the formation of a viscous string of more than 5 mm; and b) hvKp - all cases with positive string test with the presence of either KI or K2 gene [4, 8]. This study was approved by the Medical Research and Ethic Committee, Ministry of Health Malaysia (Reference Number: NMRR-20-3146-57787 (IIR)) and from the Universiti Sains Malaysia (Reference number: USM/JEPeM/21020183).

### Identification of hmKp and hvKp isolates

Specimen growth for *K. pneumoniae* isolates were proceeded for further bacterial identification based on colony morphology, gram-staining, biochemical tests, Vitek2 Gram-negative (GN) identification card (BioMerieux, Marcy Ietoile, France) and/or MALDI-TOF (Bruker Daltonik, Billerica, Massachusetts, USA). Once the colonies were identified as *K. pneumoniae*, further molecular confirmation for housekeeping gene (*gapA*) were performed. All 180 isolates of K*. pneumoniae* were identified for their hypermucoviscosity characteristic by the string test method while detection of capsular genes (K1 and K2) was carried out by PCR methods. The presence of other virulence genes (*rmpA*, *rmpA2*, *aerobactin*, *peg-344* and *magA*) were also analyzed by PCR method to investigate their association with the capsular genes.

### Preparation of bacterial DNA template

Bacterial DNA template for all 180 isolates of *K. pneumoniae* was prepared by suspending a loopful of colonies from overnight culture plate in 1.5 ml tube containing 400 µL of DNase-free distilled water. The suspension was boiled for ten minutes at 100°C and centrifuged at 13, 000 × *g* for 5 minutes to remove the cell debris. The supernatant was transferred into a new tube and used as DNA template in PCR reaction.

### Multiplex PCR amplification of the target genes

Two separate multiplex PCR assays were developed to target the housekeeping gene of *K. pneumoniae* (*gapA*) as an internal control, virulence-associated genes (*rmpA*, *rmpA2*, *iucA*, *magA*, and *peg3*), and the two most common hypervirulent *K. pneumoniae* capsular serotype genes (K1 and K2). Assay 1 was developed to target the K1, *magA* and *peg-344* genes in one system, while Assay 2 targeted the K2, *gapA*, *rmpA*, *rmpA2*, and *iucA* genes. The primers used are listed in the Table 1.

**Table 1.**
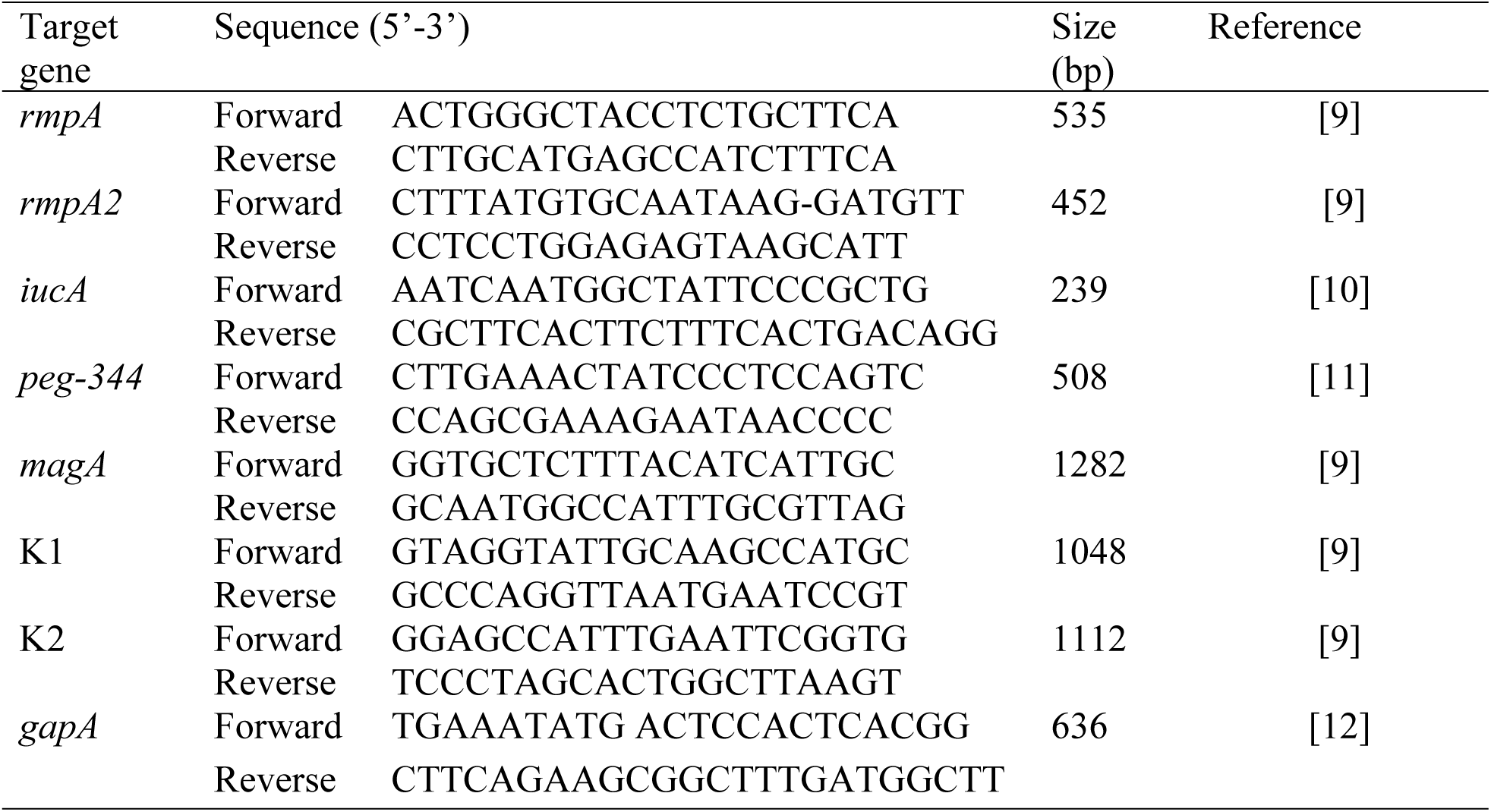
Primers used for the optimization of multiplex PCR assay.

For each Assay 1 and 2, a total volume of 20 µl per reaction was prepared, containing a set of forward and reverse primers (final concentration ranged between 0.125 and 1 pmol/μl), *Taq* DNA polymerase (1 unit/μl), My*Taq* Red Mix buffer solution (1.25×) (Bioline Reagents Ltd., UK), DNA template and molecular grade PCR water. The PCR was performed using a Mastercycler Gradient (Eppendorf, Hamburg, Germany) with an initial denaturation at 95^◦^C for 5 min, 30 cycles consisting of denaturation at 95^◦^C for 30 sec, annealing for 30 sec at 55^◦^C and extension at 68◦C for 1 min, followed by a final extension at 72^◦^C for 5 min. The PCR products were electrophoresed through 2% agarose gel (Promega, Madison, USA) at 80 volts for 75 min. Each DNA sample was subjected to both Assays 1 and 2 of the developed multiplex PCR to determine the distribution of virulence genes in all 180 *K. pneumoniae* isolates.

### Record tracing and data collection

Medical records of the patients with confirmed hvKp infections were retrieved and reviewed. Patients’ data, including demographic, clinical manifestations, underlying medical illnesses and clinical diagnosis were recorded.

### Statistical analysis

Statistical analysis was performed using SPSS software version 26 (SPSS Inc., Illinois, USA). The demographic and clinical presentations of patients were analyzed by descriptive analysis. Categorical data was presented as frequency and percentage, while numerical data was presented as mean and standard deviation (SD). The Chi Square Test and Simple Logistic Regression (SLR) tests were done for univariate and multivariate analyses. A variable comparison with a p-value less than 0.05 was considered significant.

## Results

### Clinical and microbiological characteristics of hvKp and non-hvKp isolates

Of the total 180 *K. pneumoniae* isolated from various types of specimens from four main hospitals in Kelantan, common cases were found in tracheal aspirate (27.8%, n = 50), blood (24.4%, n = 44), and sputum (21.7%, n = 39) (Figure 1). A positive string test was identified in 43 (23.9%) isolates. Of this number, only 17 (9.4%) isolates were detected to have hvKp characteristics. As described earlier, hvKp is defined in this study as *K. pneumoniae* isolates that exhibit a positive string test in the presence of either the KI or K2 genes.

**Figure 1.**
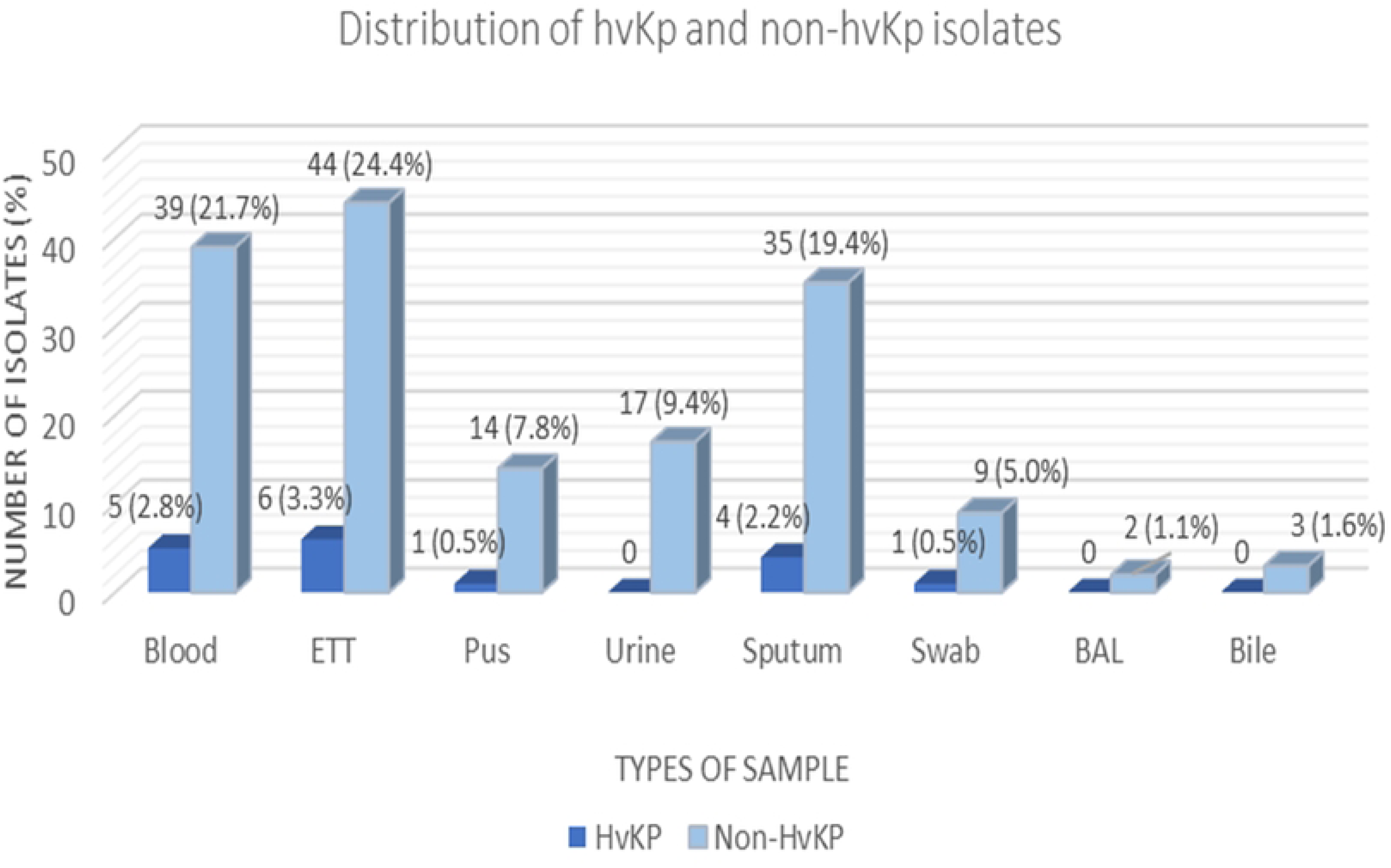
Distribution of hvKp (n = 17) and non-hvKp (n = 163) isolates based on the types of samples (n = 180).

### Social-demographic data and clinical presentations of patients infected with hvKp

The demographic data of patients infected with hvKp were reviewed and described in Table 2. The age of the patients ranged from one to 85, with a mean of 49.63 years. Patients aged less than 60 years (58.8%) were found to acquire more hvKp infections than those above 60 years. Additionally, male patients had higher infection cases than females. Respiratory infections were found to be the most common clinical presentation of hvKp infection in our study.

**Table 2:** Social-demographic data and clinical presentations of patients infected with hypervirulent *K. pneumoniae* (hvKp) (n = 17).

| **Variable** | **Options** | **Frequency**<br>**(n = 17)** | **%** |
| --- | --- | --- | --- |
| **Ages** | >60 years | 7 | 41.2 |
|  | ≤60 years | 10 | 58.8 |
| **Gender** | Male | 13 | 76.5 |
|  | Female | 4 | 23.5 |
| **Clinical**<br>**Presentation** | Pneumonia | 12 | 70.5 |
|  | Abdominal pathology | 0 | 0 |
|  | Liver abscess | 1 | 5.9 |
|  | Sepsis | 2 | 11.8 |
|  | Others (e.g: malignancy, soft tissue infection) | 2 | 11.8 |
| **Comorbid** | Diabetes mellitus | 2 | 11.8 |
|  | Hypertension | 6 | 35.3 |
|  | Pulmonary disease | 3 | 17.6 |
|  | Chronic kidney disease | 0 | 0 |
|  | Cardiovascular disease | 1 | 5.9 |
|  | No comorbidities | 5 | 29.4 |
| **Types of samples** | Blood | 5 | 29.4 |
|  | Endotracheal tube | 6 | 35.3 |
|  | Sputum | 4 | 23.5 |
|  | Pus | 1 | 5.9 |
|  | Swab | 1 | 5.9 |
| **Outcome** | Discharged | 15 | 88.2 |
|  | Death | 2 | 11.8 |

### Molecular characteristics of hvKp and non-hvKp isolates

The hvKp and non-hvKp isolates were further characterized using molecular assays based on the presence of virulence genes. The developed multiplex PCR assays simultaneously targeted K2, *gapA*, *rmpA*, *rmpA2* and *iucA* genes in Assay 1; and K1, *magA* and *peg-344* in Assay 2 (Figure 2).

**Figure 2.**
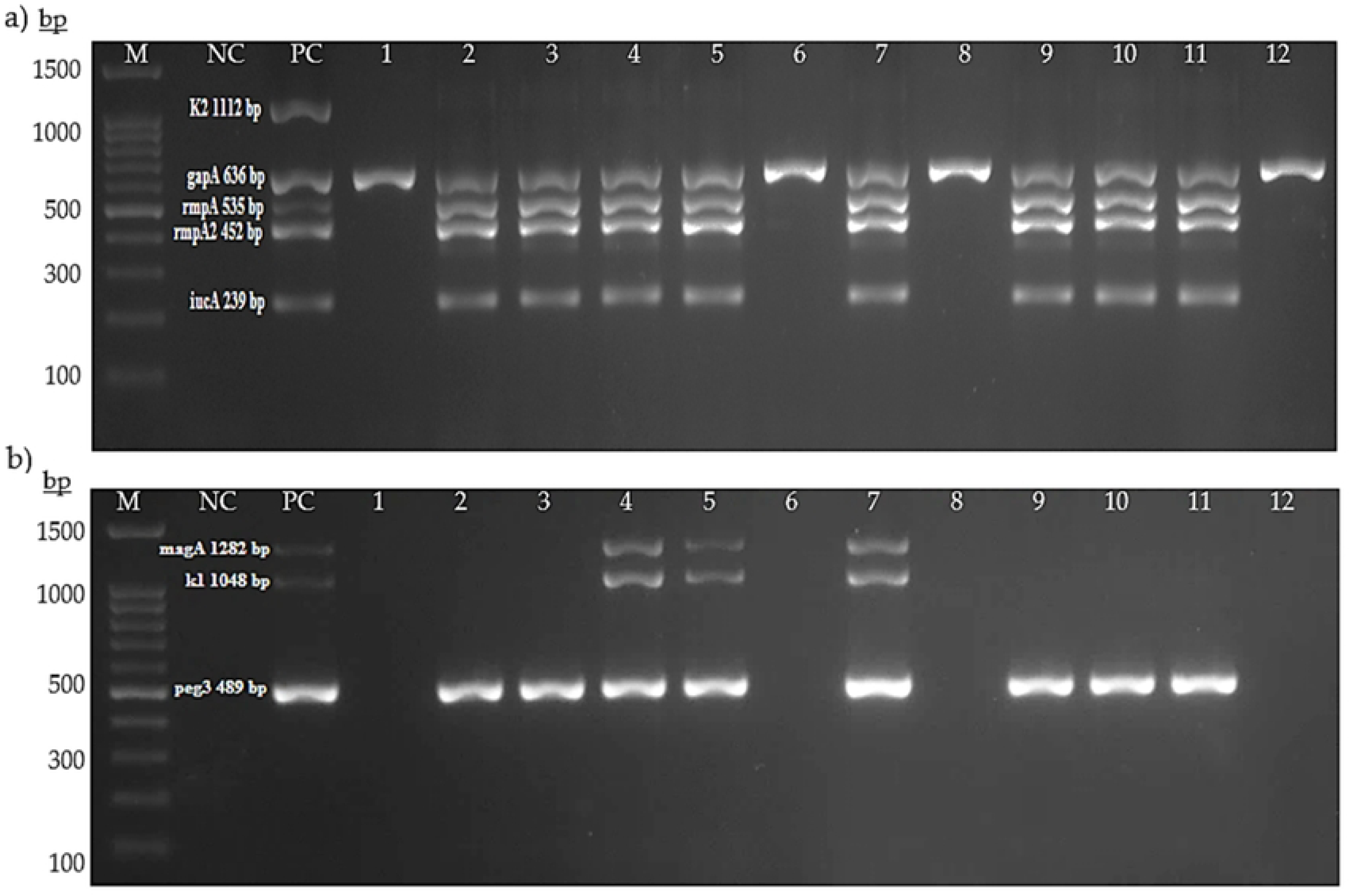
Representative image of agarose gel electrophoresis of multiplex PCR Assay 1 (a) and Assay 2 (b). Assay 1 and Assay 2 targeted K2 serotype (1112 bp), *gapA* (636 bp), *rmpA* (535 bp), *rmpA2* (452 bp) and *iucA* (239 bp); and K1 serotype (1048 bp), *magA* (1282 bp), and *peg-344* (636 bp), respectively. Lane M: 1000plus bp DNA ladder, lane NC: negative control, lane PC: positive control, lanes 1 - 12: *K. pneumoniae* isolates.

Among the total 180 *K. pneumoniae* isolates, K1 and K2 capsular serotypes were detected in 11.1% (n = 20/180) and 6.1% (n = 11/180), respectively. In contrast, 82.8% (n = 149/180) of the isolates lacked the K1/K2 capsular serotype-specific (*cps*) genes. This group was classified as non-K1/K2. In further analysis, only 50% (n = 10/20) and 63.6% (n = 7/11) of the K1 and K2 isolates expressed hypermucoviscous characteristics by string test positive, respectively. The K1 serotype exhibited 100% (20/20) positive detection for *magA, iucA* and *rmpA2* virulence genes. High detection rates were also observed for *rmpA* (95%, n = 19/20) and *peg-344* genes (80%, n = 16/20). Meanwhile, a majority of the K2 serotypes in this study were found to be positive for *iucA* (91%, n = 10/11), *rmpA* (91%, n = 10/11), *rmpA2* (82%, n = 9/11), and *peg-344* (82%, n = 9/11). None of the K2 serotypes were positive for the *magA* virulence gene.

### Distribution of virulence genes associated with hypermucoviscous ***K. pneumoniae* (hmKp) and non-hypermucoviscous *K. pneumoniae*** (non-hmKp)

Apart from the characteristic of hvKp, a total of 43 (23.9%) cases of positive string tests (hmKp) were recorded. Of these 43 hmKp isolates, 23.3% (n = 10/43) and 16.3% (n = 7/43) were K1 and K2 serotypes, respectively (Figure 3). Overall, the *rmpA* (81.4%, n = 35/43) gene was found to be the most prevalent in the hmKp strains, followed by the *iucA* (69.8%, n = 30/43) and *peg-344* (69.8%, n = 30/43), *rmpA2* (65.1%, n = 28/43) and *magA* (23.3%, n = 10/43) genes.

**Figure 3.**
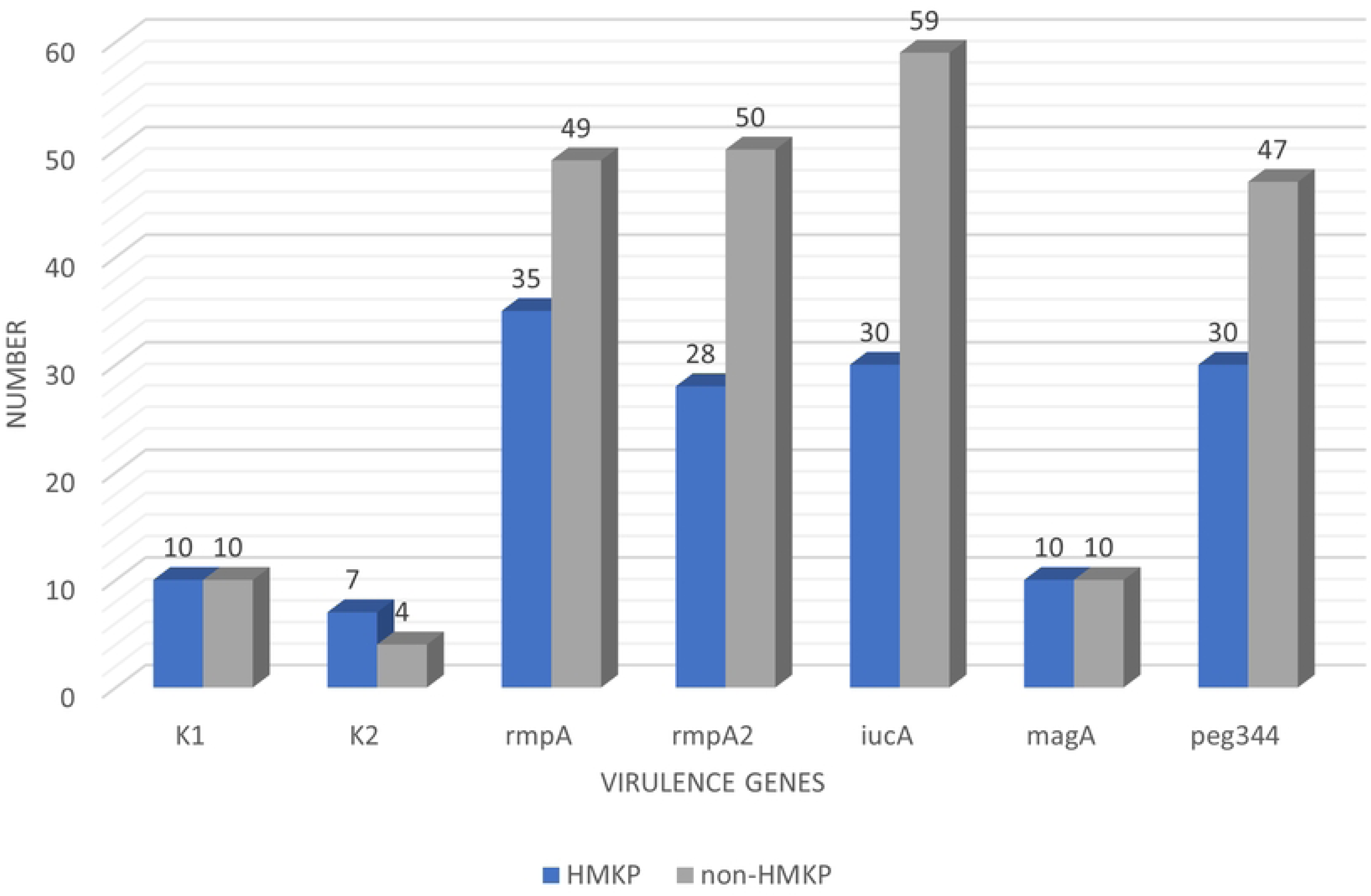
Distribution of virulence genes associated with hmKp (string test positive, n = 43) and non-hmKp (string test negative, n = 137).

### Distribution of virulence genes associated with K1 and K2 capsular serotypes

All virulence genes (*rmpA*, *rmpA2*, *iucA*, *magA*, and *peg-344*) were statistically significantly associated with K1 (p<0.001). A significant association between *rmpA*, *rmpA2*, *iucA,* and *peg-344* with K2 (p<0.05) was also observed (Table 3). Apart from that, *rmpA*, *rmpA2*, *iucA*, *magA*, and *peg-344* were significantly associated with hypermucoviscosity (p<0.005).

**Table 3.**
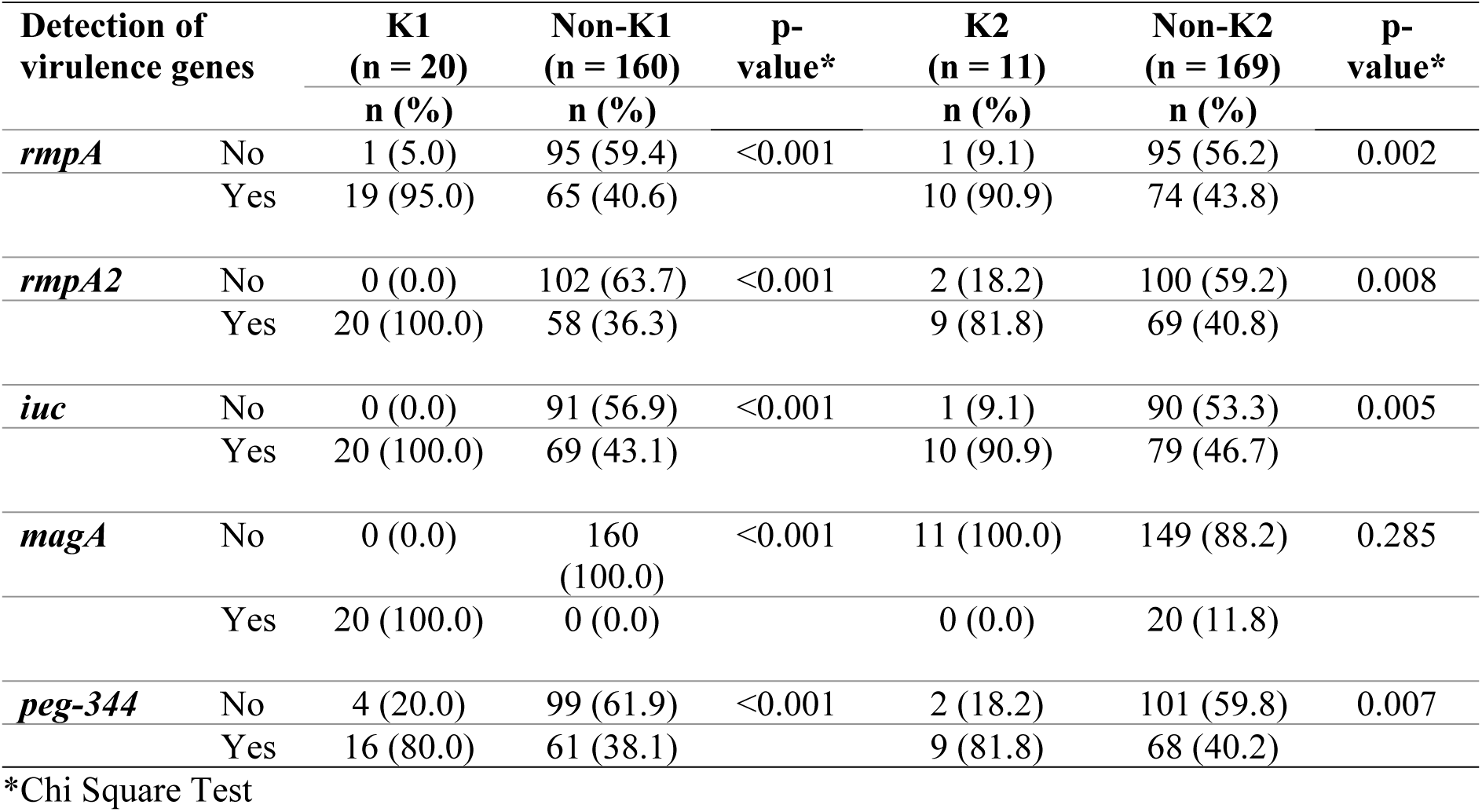
Association of the virulence-associated genes with capsular serotype K1 and K2.

## Discussion

Until today, the terminology of “hypervirulent” *K. pneumoniae* remains controversial and varies among studies, as there is still no consensus on the definition of hvKp. Even though it has been suggested that host, pathogen, and host-pathogen interactions should be considered comprehensively for defining hvKp [9] most of the studies focused on the pathogen alone. Early reports used the hypermucoviscous phenotype to define hypervirulent strains [13, 14]. However, consequent *in vitro* and *in vivo* analyses by the later studies have revealed that hypermucoviscosity alone is insufficient to describe hvKp because: i) there are disagreements between phenotypic hypermucoviscous and genotypic virulence characteristics [15, 16]; and ii) the string test has been shown to have low sensitivity and specificity ^(1, 10)^. Hence, in parallel with hypermucoviscosity, recent studies used several virulence factors, such as K1, K2, aerobactin, and *rmpA* genes, as the more reliable characteristics to describe hvKp [9, 17].

In this study, hvKp is specifically defined when the *K. pneumoniae* strains possess both of these criteria: i) hypermucoviscosity; and ii) positive detection of K1 or K2. The similar definition of hvKp was also used by a previous study conducted in Japan [4]. Based on these criteria, this study identified 23.9% (n = 43) hmKp and 9.4% (n = 17) hvKp among 180 *K. pneumoniae* isolates. In a retrospective study conducted at a single centre in Malaysia, the proportion of hmKp was 7.5% among 120 carbapenem-resistant *K. pneumoniae.* The majority of the isolates were from hospital-acquired or healthcare-associated infections, and none of the hmKp strains harboured K1 or K2 serotypes [18]. Meanwhile, in China, 33% of the isolates from hospitalized patients were characterized as hmKp (19). In endemic areas, the prevalence of hvKp among *K. pneumoniae* is high, ranging from 12% to 45% [2]. A retrospective genomic epidemiology study reported a higher prevalence of community-acquired “hypervirulent” strains in South and Southeast Asia. However, this study solely characterized hvKp strains using genome sequencing and *K. pneumoniae*-specific genomic typing tools for the presence of hypervirulence-associated loci: K1 and K2 capsular serotypes (18%), yersiniabactin (49%), aerobactin (28%)*, rmpA* (18%)*, rmpA2* (16%), and *peg-344* (19%), in 331 *K. pneumoniae* BSI isolates.

With regard to virulence factors, all the genes investigated in this study — *rmpA*, *rmpA2*, *iucA, magA*, and *peg-344*, including the capsular serotype antigens K1 and K2, were significantly associated with hypermucoviscosity (p<0.05). Interestingly, the *rmpA*, *rmpA2*, *iucA,* and *peg-344* genes were detected in all 17 hvKp isolates, which shows that these genes are highly expressed in hvKp. The proportions of K1 and K2 capsular serotypes among 180 isolates were 11.1% (n = 20) and 6.1% (n = 11), respectively, which were found to be lower than the other tested virulence genes. Although there are presently more than 70 reported K-serotypes, capsular serotypes K1 and K2 have frequently resulted in severe infections [9]. Besides, K1 and K2 are the dominant capsule serotypes that are strongly related to hypervirulent strains [10, 14]. K1 is the most predominant serotype in Asia and was reported to be highly associated with hvKp infections compared to K2 [20].

The associations of virulence genes *rmpA*, *rmpA2*, *iucA*, and *peg-344* with K1 and K2 serotypes were found to be statistically significant (p<0.05). Aerobactin (*iucA*) has been used as a molecular biomarker for hvKp [17]. This gene was present in 70% (n = 30) of the hmKp pathotype in this study and was the most common virulence factor detected (49.4%, n = 89) among all isolates. Hence, the use of aerobactin as a sole biomarker for hvKp detection is unlikely. The *magA* gene is significantly associated with K1 (p<0.05) because this gene was claimed to be a serotype-specific gene for K1 [21], justifying its absence in K2 serotypes. Based on the significant associations of the virulence genes with hypermucoviscosity and K1/K2 capsular serotypes, the simultaneous detection of *rmpA*, *rmpA2*, *magA* (for K1 serotype)*, iucA*, and *peg-344* genes in one single hmKP isolate could represent accurate genetic markers for defining a hvKp strain in the future.

Alongside the pathogen, this study also investigated the characteristics of the host (patients). Demographic data of the 17 cases infected by hvKp showed that more than half of the patients were under 60 years old. The finding was consistent with a previous study reporting that patients with hvKp tend to be younger and otherwise healthy individuals but present with severe disease [7]. Specific gender was not a recognized risk factor; however, in this study, the majority of the patients infected with hvKp were male (76.5%, n = 13). The similar finding was also reported by another study, in which men are more likely to be infected than women [22]. In terms of clinical manifestations, the patients infected with hvKp were mostly diagnosed with pneumonia (70.6%, n = 12). Patients who presented with bacteremic community-acquired pneumonia were increasingly being reported in the Asia-Pacific region and South Africa [3, 23]. Respiratory infection was reported as the second most common after liver abscess [22]. However, only one single case of liver abscess was found in this study among the hvKp isolates.

This discrepancy may be due to geographical differences in exposure and circulating strains. Hypertension was the highest underlying comorbidity (35.5%), followed by diabetes mellitus (11.8%). Some studies suggested that the connection between diabetes mellitus and hvKp could vary by region or clinical presentation [3]. About 88% of hvKp infected-patients were eventually discharged, with the mortality rate for hvKp infection was 12% (n = 2). This was slightly lower than the previous retrospective study reported in Beijing, China [24].

The study’s strength includes the involvement of four major hospitals, which could reflect the situation in Kelantan state. However, the one-year duration of the study limits the sample collection. Likewise, adding more virulence and capsular serotype genes for hvKp, such as serotypes K5, K20, K54, and K57, may expand the recovery of hypervirulent strains, although their prevalence among hvKp is low [13]. Our study also lacks information on the antimicrobial susceptibility profiles of the *K. pneumoniae* isolates. Hence, antibiogram data should be incorporated into hvKp surveillance studies in the future, as multidrug-resistant hvKp strains are increasingly being reported worldwide [25, 26].

## Conclusion

In conclusion, based on the positive detection of hmKp and K1 or K2 serotypes, the prevalence of hvKp in this study was found to be 9.4%. HvKp infections were more common in men and individuals with hypertension. Pneumonia was the most common clinical diagnosis in hvKp-infected patients. The mortality rate for hvKp infection was 12%. Molecular detection of virulence factors is more appropriate as using the string test alone for hvKp detection could be misleading. The presence of all biomarkers (*rmpA*, *rmpA2*, *magA* (for K1 serotype)*, iucA*, and *peg-344*) in hmKp, along with clinical presentations, could accurately define hvKp. This is important for effective clinical management and epidemiological surveillance.

## Data Availability

All relevant data are within the manuscript and its Supporting Information files.

## Acknowledgement

The authors would like to thank the Department of Medical Microbiology and Parasitology, Universiti Sains Malaysia; Hospital Universiti Sains Malaysia; Hospital Raja Perempuan Zainab II; Hospital Sultan Ismail Petra; and Hospital Tanah Merah for providing the bacterial strains and granting us permission to review patients’ records.

## References

1. Catalan-Najera JC, Garza-Ramos U, Barrios-Camacho H. Hypervirulence and hypermucoviscosity: Two different but complementary *Klebsiella* spp. phenotypes? Virulence. 2017;8(7):1111–23.

2. Choby JE, Howard-Anderson J, Weiss DS. Hypervirulent Klebsiella pneumoniae - clinical and molecular perspectives. J Intern Med. 2020;287(3):283–300.

3. Russo TA, Marr CM. Hypervirulent *Klebsiella pneumoniae*. Clin Microbiol Rev. 2019;32(3).

4. Ikeda M, Mizoguchi M, Oshida Y, Tatsuno K, Saito R, Okazaki M, et al. Clinical and microbiological characteristics and occurrence of *Klebsiella pneumoniae* infection in Japan. Int J Gen Med. 2018;11:293–9.

5. Zhang Y, Zhao C, Wang Q, Wang X, Chen H, Li H, et al. High Prevalence of Hypervirulent *Klebsiella pneumoniae* Infection in China: Geographic Distribution, Clinical Characteristics, and Antimicrobial Resistance. Antimicrob Agents Chemother. 2016;60(10):6115–20.

6. Casanova C, Lorente JA, Carrillo F, Perez-Rodriguez E, Nunez N. Klebsiella pneumoniae liver abscess associated with septic endophthalmitis. Arch Intern Med. 1989;149(6):1467.

7. Marr CM, Russo TA. Hypervirulent *Klebsiella pneumoniae*: a new public health threat. Expert Rev Anti Infect Ther. 2019;17(2):71–3.

8. Shon AS, Bajwa RP, Russo TA. Hypervirulent (hypermucoviscous) Klebsiella pneumoniae: a new and dangerous breed. Virulence. 2013;4(2):107–18.

9. Liu C, Guo J. Hypervirulent *Klebsiella pneumoniae* (hypermucoviscous and aerobactin positive) infection over 6 years in the elderly in China: antimicrobial resistance patterns, molecular epidemiology and risk factor. Ann Clin Microbiol Antimicrob. 2019;18(1):4.

10. Sanikhani R, Moeinirad M, Shahcheraghi F, Lari A, Fereshteh S, Sepehr A, et al. Molecular epidemiology of hypervirulent *Klebsiella pneumoniae*: a systematic review and meta-analysis. Iran J Microbiol. 2021;13(3):257–65.

11. Russo TA, Olson R, Fang CT, Stoesser N, Miller M, MacDonald U, et al. Identification of Biomarkers for Differentiation of Hypervirulent *Klebsiella pneumoniae* from Classical K. pneumoniae. J Clin Microbiol. 2018;56(9).

12. Diancourt L, Passet V, Verhoef J, Grimont PA, Brisse S. Multilocus sequence typing of *Klebsiella pneumoniae* nosocomial isolates. J Clin Microbiol. 2005;43(8):4178–82.

13. Lin ZW, Zheng JX, Bai B, Xu GJ, Lin FJ, Chen Z, et al. Characteristics of Hypervirulent *Klebsiella pneumoniae*: Does Low Expression of rmpA Contribute to the Absence of Hypervirulence? Front Microbiol. 2020;11:436.

14. Liu YM, Li BB, Zhang YY, Zhang W, Shen H, Li H, et al. Clinical and molecular characteristics of emerging hypervirulent *Klebsiella pneumoniae* bloodstream infections in mainland China. Antimicrob Agents Chemother. 2014;58(9):5379–85.

15. Zhang Y, Zeng J, Liu W, Zhao F, Hu Z, Zhao C, et al. Emergence of a hypervirulent carbapenem-resistant *Klebsiella pneumoniae* isolate from clinical infections in China. J Infect. 2015;71(5):553–60.

16. Lin YC, Lu MC, Tang HL, Liu HC, Chen CH, Liu KS, et al. Assessment of hypermucoviscosity as a virulence factor for experimental *Klebsiella pneumoniae* infections: comparative virulence analysis with hypermucoviscosity-negative strain. BMC Microbiol. 2011;11:50.

17. Russo TA, Olson R, Macdonald U, Metzger D, Maltese LM, Drake EJ, et al. Aerobactin mediates virulence and accounts for increased siderophore production under iron-limiting conditions by hypervirulent (hypermucoviscous) *Klebsiella pneumoniae*. Infect Immun. 2014;82(6):2356–67.

18. Kong ZX, Karunakaran R, Abdul Jabar K, Ponnampalavanar S, Chong CW, Teh CSJ. The Detection of Hypermucoviscous Carbapenem-Resistant *Klebsiella pneumoniae* from a Tertiary Teaching Hospital in Malaysia and Assessment of Hypermucoviscous as Marker of Hypervirulence. Microb Drug Resist. 2021;27(10):1319–27.

19. Li W, Sun G, Yu Y, Li N, Chen M, Jin R, et al. Increasing occurrence of antimicrobial-resistant hypervirulent (hypermucoviscous) *Klebsiella pneumoniae* isolates in China. Clin Infect Dis. 2014;58(2):225–32.

20. Bilal S, Volz MS, Fiedler T, Podschun R, Schneider T. *Klebsiella pneumoniae*-induced liver abscesses, Germany. Emerg Infect Dis. 2014;20(11):1939–40.

21. Guo Y, Wang S, Zhan L, Jin Y, Duan J, Hao Z, et al. Microbiological and Clinical Characteristics of Hypermucoviscous *Klebsiella pneumoniae* Isolates Associated with Invasive Infections in China. Front Cell Infect Microbiol. 2017;7:24.

22. Siu LK, Yeh KM, Lin JC, Fung CP, Chang FY. *Klebsiella pneumoniae* liver abscess: a new invasive syndrome. Lancet Infect Dis. 2012;12(11):881–7.

23. Wu H, Li D, Zhou H, Sun Y, Guo L, Shen D. Bacteremia and other body site infection caused by hypervirulent and classic *Klebsiella pneumoniae*. Microb Pathog. 2017;104:254–62.

24. Li J, Ren J, Wang W, Wang G, Gu G, Wu X, et al. Risk factors and clinical outcomes of hypervirulent *Klebsiella pneumoniae* induced bloodstream infections. Eur J Clin Microbiol Infect Dis. 2018;37(4):679–89.

25. Hao M, Shi X, Lv J, Niu S, Cheng S, Du H, et al. In vitro Activity of Apramycin Against Carbapenem-Resistant and Hypervirulent *Klebsiella pneumoniae* Isolates. Front Microbiol. 2020;11:425.

26. Chen J, Zhang H, Liao X. Hypervirulent *Klebsiella pneumoniae*. Infect Drug Resist. 2023;16:5243–9.

